# Implementation, Indications, and Rationale for Modified Free Water Protocols in Paediatric Dysphagia: A UK Survey

**DOI:** 10.1101/2025.11.18.25340471

**Authors:** Alanna Thompson, Jessica De Bolfo

## Abstract

**Background:** Modified Free Water Protocols (MFWPs) are increasingly referenced in UK paediatric dysphagia practice despite no published paediatric evidence of safety or efficacy.

**Aims:** To examine how, for whom, and why MFWPs are used or discussed by UK Speech and Language Therapists (SLTs), whether clinicians can reference any supporting evidence, and how outcomes are monitored.

**Methods:** Cross-sectional online survey of UK paediatric dysphagia SLTs with Likert-scale, yes/no, and open-ended items; data analysed descriptively and thematically.

**Results:** Sixty-eight clinicians responded; 64 % (44/68) reported using an MFWP, yet only 12 % (8/68) had local written guidance. Most adaptations derived from adult FWPs, most often the Frazier protocol. No respondent cited paediatric evidence or justification for “cooled-boiled/’sterile’” water. MFWPs were applied to children with thin-fluid aspiration, refusal of thickeners, chronic respiratory disease, and severe neurological impairment; groups that would not meet adult FWP candidacy. Outcome monitoring centred on respiratory health but without details of how or when monitoring occurred; hydration and functional indices were rarely reported.

**Conclusions:** UK clinicians are applying heterogeneous, unvalidated adaptations of adult FWPs to children. The use of “cooled-boiled/ “sterile’” water is unsupported and distracts from evidence-based safeguards such as oral care and supervision. National guidance and paediatric outcome evidence are urgently required.

## Introduction

Aspiration of thin fluids (IDDSI Level 0) in children with dysphagia is clinically significant and associated with pneumonia, dehydration, and reduced quality of life.^1^–^3^ Silent aspiration is common and often undetected without instrumental assessment; therefore, video-fluoroscopic swallow study (VFSS) or flexible endoscopic evaluation of swallowing (FEES) are essential for accurate diagnosis and management.4-6.

Thickened fluids are frequently recommended to reduce aspiration risk, yet they may negatively affect hydration, enjoyment, and compliance.^7^–^10^ Clinicians therefore face complex trade-offs between airway protection and quality of life, particularly when children refuse thickeners or have limited access to instrumental assessment.

In adult rehabilitation, Free Water Protocols (FWPs) notably the Frazier Free Water Protocol^11^ and GF Strong Water Protocol^15^ permit carefully selected patients access to water under strict safeguards: medical stability, adequate cognition, absence of active pneumonia, meticulous oral hygiene, water only between meals, supervision, and medication precautions. In adults, when applied to appropriate candidates, pneumonia incidence does not increase.^12^–^14^,^16^,^17^ Importantly, adult FWPs presume potable water and do not require “cooled-boiled” or “sterile” water.

Within paediatric practice, references to so-called Modified Free Water Protocols (MFWPs) have emerged, yet no paediatric data confirm their safety or efficacy. Reported approaches often mirror adult FWPs but incorporate untested modifications, most notably the recommendation for “cooled-boiled” or “sterile” water.

This survey was intended to examine how, for whom, and why MFWPs are implemented in UK paediatric practice; to determine whether clinicians possess or can reference any evidence supporting the paediatric use of MFWPs, including the “cooled-boiled/’sterile’” modification; and to describe how outcomes are monitored.

### Caveat regarding the term “protocol”

Although the phrase “modified free water protocol” (MFWP) is commonly used in UK paediatric dysphagia practice, it does not accurately reflect a validated, standardised, or evidence-based protocol. The term “protocol” implies a formally developed and consistently applied intervention with defined eligibility criteria, safeguards, implementation procedures, and outcome measures, which is not yet established for paediatric populations. Nevertheless, for the purposes of reporting and interpreting survey findings, the term “MFWP” will be used in this paper to reflect the colloquial terminology currently used by clinicians to describe the practice of allowing children with known or suspected aspiration of fluids to consume water orally under specified conditions or restrictions. Findings however challenge the appropriateness of this terminology and highlight the need for more accurate, specific language.

## Methods

### Design and Participants

A descriptive, cross-sectional online survey was distributed between [insert months/year] to UK-based HCPC-registered SLTs managing paediatric dysphagia. Recruitment used national and regional networks (Paediatric Clinical Excellence Network, acute paediatric SLT groups). Participation was voluntary and anonymous.

### Survey Instrument

The questionnaire comprised Likert-scale, yes/no, and open-ended items exploring: demographics and setting; MFWP use vs non-use; confidence and perceived evidence; patient selection and contraindications; implementation details (timing, oral care, supervision, and recommendations regarding “cooled-boiled/’sterile’” water); protocol origin and modification (adult FWPs cited); outcome monitoring and service integration; and an explicit request for citations supporting paediatric MFWPs or specific modifications. Five senior SLTs reviewed the draft for clarity and face validity.

### Data Analysis

Only responses from UK-based clinicians were analysed to ensure contextual comparability and consistency with national policy frameworks. The survey incidentally reached a small number of international respondents (n = 11; Australia n = 6, New Zealand n = 5), whose data were excluded from analysis to preserve focus and interpretive validity. Quantitative items were summarised as frequencies and percentages using Microsoft Excel 365. Qualitative responses underwent inductive thematic analysis by both authors to identify patterns in how, for whom, and why MFWPs were used. Keyword searches tallied adult protocols named (“Frazier”, “GF Strong”), references to “boiled/cooled/sterile”, and descriptions of outcome monitoring.

### Governance and Ethics

St George’s Hospital Research & Information Governance confirmed that NHS Research Ethics Committee review was not required (service evaluation/professional practice study). No patient-identifiable data were collected.

## Results

UK respondents n = 68

### Geographical and Professional Profile

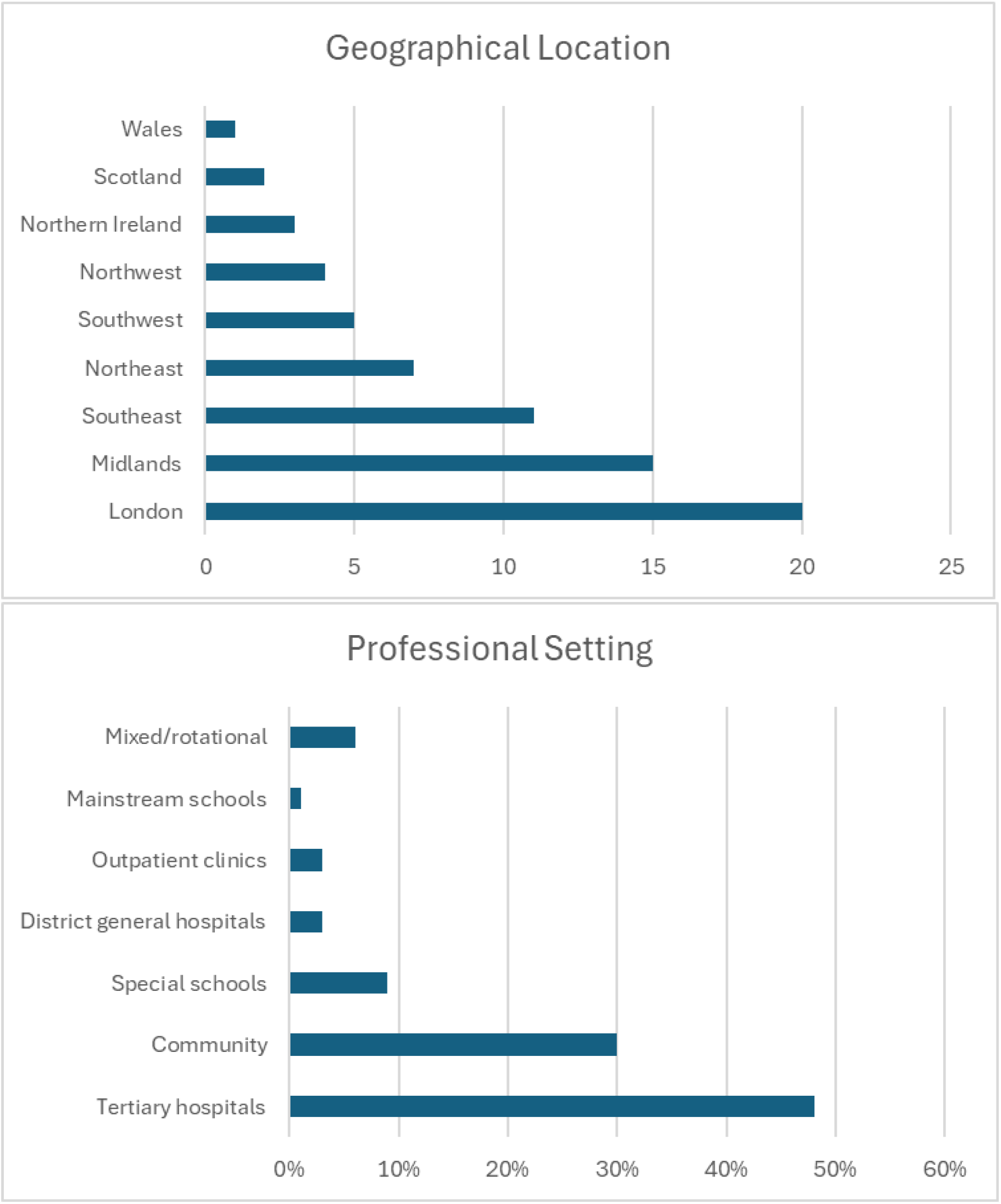

### Self-reported use, confidence levels, and guidance for MFWP use

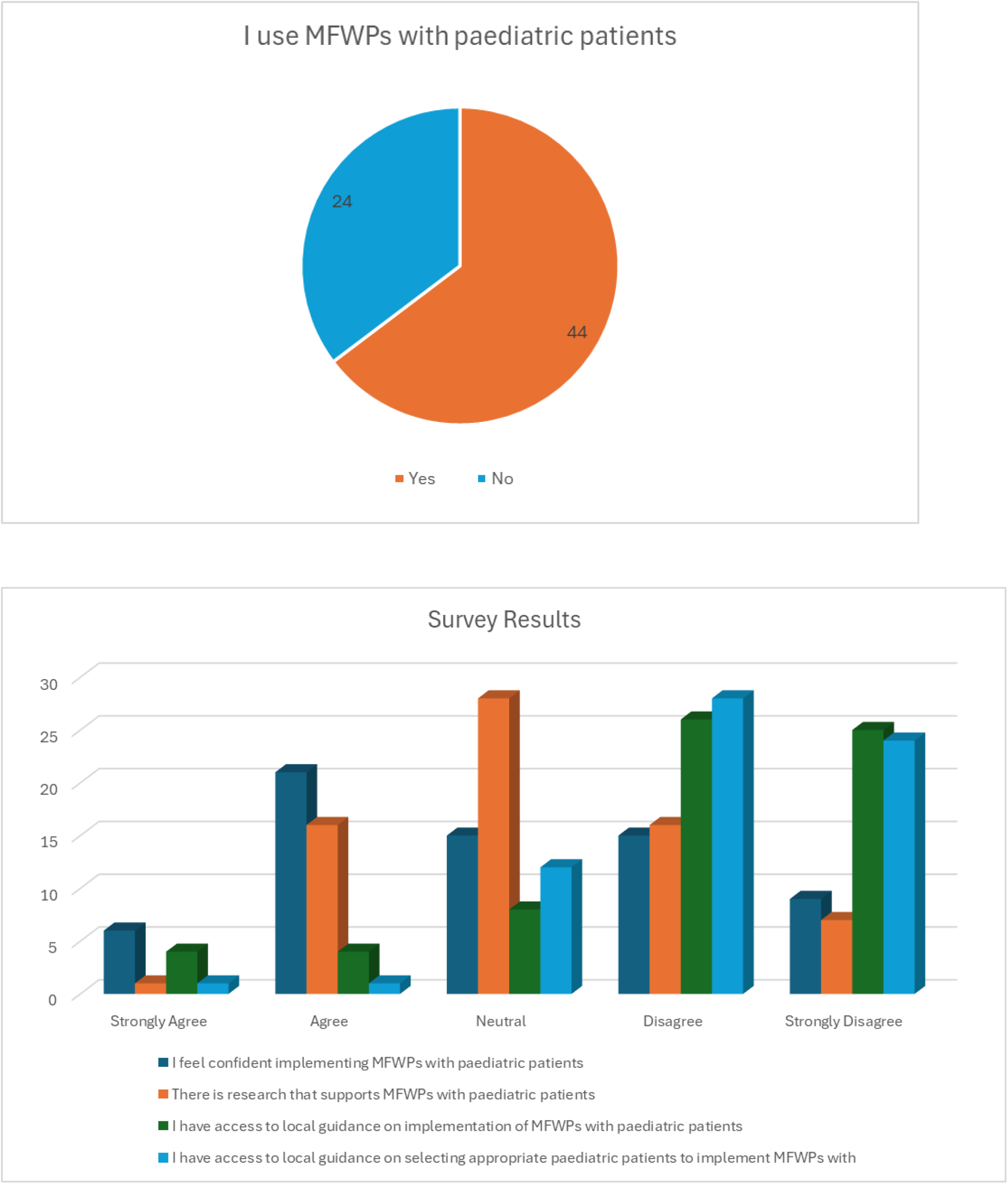

### Adult Protocols and Modifications

Most clinicians reported adapting an adult FWP, but did not specifically cite the adult FWP that they had modify. The Fraizer FWP ^11^ was cited 10 times. No other FWP was cited. Interestingly, although the FFWP emphasises oral hygiene, between-meals timing, supervision, and medication rules; it does not include a recommendation for “cooled-boiled/’sterile’” water.^11^–^17^.

### “Cooled-boiled/ “Sterile” Water

Mentions of “boiled/cooled” appeared in at least 8 responses; “sterile” in 4. No participant cited supporting evidence for this. It should be noted that true sterility requires aseptic preparation, handling, and storage, and no respondant described sterile preperation, storage, and handling of “sterile” water. Additionally, no respondant provided information on how they instruct families to sterilise water, with many respondants referring to sterile and cooled boiled water interchangably.

WHO and CDC guidance indicate that bringing water to a rolling boil for one minute inactivates pathogens where tap water is unsafe,^18^–^21^ but in the UK drinking water is already regulated to microbiological standards. Boiling therefore provides no added protection against aspiration-related infection. The focus on “sterile” water risks diverting attention from the adult-FWP safeguard most relevant to pneumonia risk reduction, which is oral hygiene.

### Patient Selection and Contraindications

Indications included refusal of thickeners (at least 34), thin-fluid aspiration (at least 24), respiratory disease (at least 13), severe neurological impairment (at least 2), and community/outpatient contexts (at least 1). Many such cases would not meet adult FWP eligibility criteria (medical stability, cognition, supervision, absence of active infection).^11^–^17^ Conditions emphasised cognition (at least 11) and oral care (at least 4). Contraindications included active chest infection, respiratory compromise, overt cough, and severe neurological impairment. Timing and supervision were infrequently described.

### Outcome Monitoring

Monitoring practices were highly variable and often vaguely described. Most respondents referred to “respiratory health” or “chest infections,” but none explained how or when these outcomes were monitored, whether via clinical review, caregiver report, or record audit. No consistent timeframe, metric, or cessation criterion was reported. Hydration indices (for example, fluid charts) were mentioned by at least four respondents and caregiver feedback by a similar number; growth monitoring was rarely mentioned.

## Discussion

Collectively, findings show that MFWPs are being used in paediatric dysphagia practice despite no paediatric evidence. Clinicians appear motivated by hydration and quality-of-life concerns, particularly where thickeners are refused or instrumental assessment is delayed. Yet fundamental safeguards central to adult FWP, including oral hygiene, between-meals timing, supervision, and medication management were inconsistently reported, while unvalidated additions (notably “cooled-boiled “sterile’” water) have appeared without evidential rationale.

The introduction of “cooled-boiled/’sterile’” water has no published origin and misuses the term sterile. WHO and CDC guidance on boiling water pertains to unsafe public water supplies, not aspiration prevention; UK tap water is already safe to drink.^18^–^21^ Using the terms “cooled boiled” and “sterile” water interchangably is erroneous. True sterility requires sterile preperation, handling, and storage. Therefore, focus on water sterility is therefore moot in this context and potentially distracting. Aspiration pneumonia is primarily associated with oral bacterial load and biofilm,^22^–^24^ supporting the adult-FWP focus on oral care as a risk-mitigation measure.

Reported patient groups often included children with severe neurological or respiratory compromise individuals explicitly excluded from adult FWPs. Referring to such discretionary local practices as “protocols” risks conveying an illusion of validation and may mislead families and colleagues regarding the evidential status of these interventions.

Although many clinicians stated that respiratory outcomes were monitored, none described how these were tracked, whether prospectively, by audit, or opportunistically during follow-up. The absence of defined frequency, metrics, or triggers for cessation underscores the lack of standardisation and precludes reliable evaluation of safety or efficacy.

### Limitations

Interpretation should consider limitations. The sample was modest and self-selected; data were self-reported; free-text coding may under-represent themes. No patient outcomes were measured, so causation cannot be inferred. Despite these constraints, the consistency of themes across diverse regions and settings suggests a genuine, system-wide evidence gap.

### Future Directions

Future research should avoid using the term “modified free water protocol” for children, as adult derived terminology and safeguards have not been validated in paediatric populations. Instead, studies should identify which child specific strategies, such as enhanced oral care, supervised access to water, timing relative to meals, hydration monitoring, and criteria for cessation, can help reduce risks associated with aspiration while supporting quality of life. Longitudinal multicentre research is recommended to evaluate safety, feasibility, hydration, respiratory outcomes, caregiver experience, and cost implications over time. Health equity analyses should examine socioeconomic and geographic disparities in access to instrumental swallow assessments and specialist dysphagia services. While this study focused on UK practice, the limited responses from Australia and New Zealand indicate emerging international interest. Future studies should therefore incorporate broader cross national scoping to understand variation in terminology, interpretation, governance, and clinical decision making across healthcare systems. National bodies should support development of consensus language and discourage use of the term protocol until paediatric specific evidence, safeguards, and standardisation exist

## Conclusions

UK paediatric SLTs are implementing MFWPs in the absence of paediatric evidence, often extending use to high-risk populations and omitting core adult-derived safeguards. The addition of “cooled-boiled/’sterile’” water is unsupported, scientifically inaccurate, and potentially distracting from oral-care and supervision measures that reduce aspiration risk. Labelling these ad-hoc approaches a “protocol” risk conveying false certainty. Clinicians should communicate the experimental nature of MFWP use to families and teams and prioritise generation of paediatric evidence.

## Data Availability

All data produced in the present study are available upon reasonable request to the authors.

## Appendix A: Survey Items, Response Formats, and Data Type

| Item No. | Survey Question | Response Format | Data Type | Notes on Construct / Domain |
| --- | --- | --- | --- | --- |
| 1 | I implement a “modified free water protocol” with selected paediatric patients | Likert scale (Strongly Agree to Strongly Disagree) | Ordinal | Assesses self-reported implementation behaviour |
| 2 | I feel confident implementing a modified free water protocol with paediatric patients | Likert scale | Ordinal | Clinical self-efficacy |
| 3 | To my knowledge, research supports the use of modified free water protocols in paediatric patients with dysphagia | Likert scale | Ordinal | Knowledge and evidence awareness |
| 4 | Modified free water protocols are associated with reduced risk compared to consumption of regular thin fluids | Likert scale | Ordinal | Risk perception and safety judgement |
| 5 | Modified free water protocols are associated with reduced risk compared to thickened fluids in paediatrics | Likert scale | Ordinal | Comparative risk judgement |
| 6 | At my workplace, we have clear guidelines regarding how to implement a modified free water protocol | Likert scale | Ordinal | Institutional governance and policy support |
| 7 | At my workplace, we have clear guidelines regarding which patients are or are not appropriate for a modified free water protocol | Likert scale | Ordinal | Governance and candidacy criteria awareness |
| 8 | Research shows modified free water protocols result in improved hydration in paediatric patients with dysphagia when compared to thickened fluids | Likert scale | Ordinal | Perceived clinical benefit (hydration) |
| 9 | In my clinical practice, parents report improved satisfaction with free water protocol compared to thickened fluids | Likert scale | Ordinal | Family reported experience |
| 10 | I have adequate understanding of the research basis for a modified free water protocol to implement it safely and confidently | Likert scale | Ordinal | Knowledge and training clarity |
| 11 | I have adequate support from the multidisciplinary team I work within to facilitate implementation of a modified free water protocol | Likert scale | Ordinal | MDT readiness / collaboration |
| 12 | Do you use any version of a modified free water protocol or cooled boiled water for children with dysphagia in your work setting? | Yes/No | Nominal | Practice adoption (binary) |
| 13 | Is the use linked with the Eating and Drinking with Acknowledged Risk (EDAR) pathway in your practice? | Yes/No | Nominal | Risk governance |
| 14 | Where do you practice? | Multiple choice (country) | Categorical | Geographic context / health system |
| 15 | What setting do you work in? | Multiple choice (clinical context) | Categorical | Clinical context / service configuration |
| 16 | Which patient groups or in which clinical situations would you recommend a free water protocol? | Open text | Qualitative | Candidacy definitions / scope |
| 17 | Are there any conditions that must be met for you to recommend a modified free water protocol? | Open text | Qualitative | Safeguards and eligibility |
| 18 | What patient conditions or factors would contraindicate the use? | Open text | Qualitative | Risk exclusion criteria |
| 19 | How do you implement a modified free water protocol or cooled boiled water in your practice? | Open text | Qualitative | Protocol adaptation and procedures |
| 20 | How do you monitor whether a modified free water protocol impacts hydration and respiratory health? | Open text | Qualitative | Outcome monitoring |
| 21 | What research evidence or existing protocols do you reference or base your recommendations on? | Open text | Qualitative | Source of influence, evidence provenance |

